# Performance of three diagnostic tests for tuberculosis among Human Immunodeficiency Virus (HIV) patients presenting to a health facility in an informal urban settlement in Nairobi, Kenya

**DOI:** 10.1101/2024.07.12.24310329

**Authors:** Eunita A. Ochola, Marshal M. Mweu

**Author notes:** These authors contributed equally to this work.

## Abstract

The absence of an accurate reference test complicates the evaluation of tuberculosis (TB) diagnostic tests among people living with Human Immunodeficiency Virus (PLWHIV). The objective of this study was to estimate (using Bayesian latent class models [BLCM]) the sensitivity (Se), specificity (Sp) and negative and positive predictive values (NPV and PPV) of sputum smear microscopy (SSM), Xpert Ultra and lipoarabinomannan antigen (LAM) tests for TB among PLWHIV in Nairobi, Kenya.

This cross-sectional study enrolled a total of 190 patients aged ≥ 18 years with presumptive TB seeking treatment at the Kibra Community Health Center Comprehensive Care Centre (CCC) clinic between September 2022 and March 2023. The diagnostic data obtained from the three tests were analysed using a BLCM framework to derive accuracy estimates of the three diagnostic tests.

The Xpert Ultra assay registered a higher Se (85.0; 95% PCI [41.4 – 99.4]) compared to LAM (26.8; 95% PCI [4.7 – 67.6]) and SSM (56.7 [16.4 – 97.4]). However, SSM had the highest Sp (99.6; 95% PCI [97.7 – 100.0]). The Xpert Ultra assay yielded the highest overall combination of Se and Sp at 80.8% (95% PCI [37.0 – 96.5]). On predictive values, SSM recorded the highest PPV at 84.5% (95% PCI [38.4 – 99.4]). Nonetheless, all the tests exhibited noticeably high NPVs (>96%).

An optimal testing approach in this low-prevalence TB setting could entail an initial screening with the more sensitive Xpert Ultra test, with any resultant positives retested with the more specific SSM test - a two-test serial testing strategy.

## Introduction

Worldwide, tuberculosis (TB) ranks among the main causes of mortality from a single pathogenic agent(1). Human Immunodeficiency Virus (HIV) infected patients have a 20 to 30 fold higher probability of contracting TB(2). As of 2017, approximately 9% of the 10 million new TB cases were people living with HIV (PLWHIV), with Africa accounting for a 72% disproportionate share(3). Kenya ranks amid the high-TB burden nations whose primary cause of death is TB(4).

In Kenya, pulmonary TB is the most prevalent type and 66% of TB cases occur among those aged ≤ 44 years(5). In line with the national TB diagnosis and treatment guidelines, the commencement of TB treatment is informed by bacteriological confirmation of *Mycobacterium tuberculosis* (MTB). However, clinical diagnosis where bacteriological confirmation is not possible may suffice for initiation of treatment(6).

Sputum smear microscopy (SSM) has been the main diagnostic tool for TB especially in developing countries (7). It is extensively used because it is easy to run, rapid, inexpensive and has high specificity (Sp) for TB diagnosis (8, 9). Nevertheless, it has a limited sensitivity (Se) of approximately 10,000 TB bacilli/ml of sputum. Notably, its poor Se poses a challenge for diagnosis of TB in PLWHIV since they have fewer bacilli in their sputum (10).

Diagnostic tests with improved Se and rapid turnaround time among PLWHIV could lead to improved patient outcomes, reduced mortality and lower progressive transmission of TB (11). The real time polymerase chain reaction (PCR) assay utilizing the gene Xpert platform such as Xpert Ultra, offers better test Se and Sp with ability to discriminate between tuberculous and non-tuberculous mycobacteria(12). Moreover, the test may detect at least 15.6 Colony forming units (CFU) of MTB/ml in sputum samples in under two hours(12). Nonetheless, the technical and operational cost implications of the diagnostic test in resource-limited settings remain a challenge(13, 14).

With sputum production among HIV infected individuals being limited(15), the need for alternative biological specimens for the diagnosis of TB arises. A rapid immunochromatographic test detecting the *Mycobacterium* cell wall antigen lipoarabinomannan (LAM) in urine samples is available as a point-of-care test for TB (16). The test is easy to run, has no need for specialized equipment or laboratory facilities and results are obtainable within 25 minutes – rendering the test field applicable. Moreover, among PLWHIV, the test displays high Sp, with Se increasing with diminishing CD4 counts(15-18).

Diagnostic studies evaluating the performance of SSM, Xpert Ultra and TB LAM among PLWHIV have previously been conducted globally (19, 20). A major shortcoming of these evaluations is that the index tests were evaluated against imperfect reference standards, thus introducing bias in the accuracy estimates of the evaluated tests(21). A superior alternative involves the use of latent class models fit using Bayesian methods that allow for concurrent evaluation of Se and Sp of two or more tests without prior knowledge of the unobserved disease status of individuals(22). Consequently, the objective here was to estimate the Se and Sp as well as the predictive values of SSM, Xpert Ultra and LAM tests for TB among HIV patients within an informal outpatient setting using a Bayesian latent class model (BLCM) framework.

## Materials and Methods

### Study setting, design and population

The study was carried out at the Kibra Community Health Centre; a level three state-run facility located in the southern part of Nairobi. Notably, the facility serves an immediate large informal-settlement population characterised by a high dual burden of HIV/AIDS and TB, which are leading causes of mortality in slums(23). It registers around150-200 out-patients daily, conducts approximately 50-60 deliveries monthly and offers comprehensive care clinic (CCC) services to over 1,500 patients per month.

A facility-based cross-sectional study was employed to evaluate the performance of the three diagnostic tests for TB. The rationale for the choice of this design stems not only from the descriptive nature of the study but also the ease of recruitment of study participants presenting to the CCC clinic for care.

### Sample size and participant selection

The required sample size was derived using the McNemar’s sample size formula for paired proportions(24):

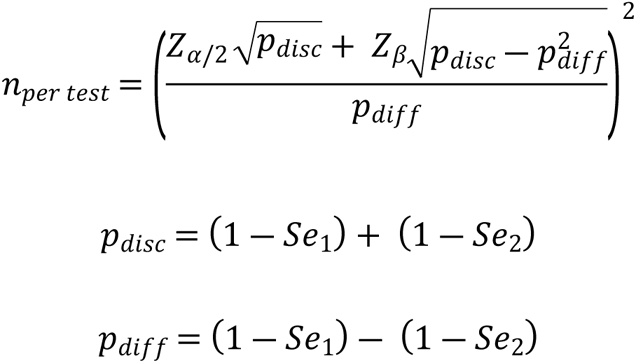

Where: 𝑛_𝑝𝑒𝑟 𝑡𝑒𝑠𝑡_= sample size required for each test, 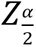(1.96) is the value required for the two-sided 95% confidence level, and 𝑍_𝛽_ (-0.84) is the value specifying the desired power of 80%. 𝑆𝑒_1_ and 𝑆𝑒_2_ are estimates of Se of the Gene xpert MTB/RIF and SSM respectively from literature i.e. 𝑆𝑒_1_is 90.3% and 𝑆𝑒_2_is 67.1% (25).

As per the above mentioned figures, a total sample size of 190 was generated, after an upward adjustment by 5% to account for non-response.

To obtain the required sample, individuals visiting the hospital’s CCC clinic were systematically random sampled in their order of arrival where every seventh person (after an initial random start) who met the eligibility criteria was selected. Eligibility was limited to PLWHIV (symptomatic or asymptomatic for TB) aged ≥18 years who at the time of recruitment (28^th^ September 2022-31^st^ March 2023) were visiting the CCC at the facility for care and consented to participation in the study. Importantly, recruitment followed triage by the clinic nurse and examination by a physician.

### Ethical considerations

Written permission to conduct this study was obtained from and approved from the Kenyatta National Hospital -University of Nairobi Ethics and Research Committee (Ref. No. KNH/ERC/R/145 and KNH/ ERC/Mod & SAE/347), the Nairobi metropolitan Services (Ref. No. EOP/NMS/HS/222), as well as the National Commission for Technology and Innovation (NACOSTI) (Ref. No. NACOSTI/P/21/14098). Moreover, a written informed consent was secured from individual patients prior to enrolling them into the study.

### Sample collection

Participants were provided with clear instructions on collection of urine and sputum samples. Of note, the LAM and SSM tests were carried out immediately at the facility’s laboratory by a trained laboratorian. Samples for the Xpert Ultra assay were stored in cooler boxes and delivered to a referral laboratory for processing. On enrolment, the participants’ sociodemographic characteristics (age, sex, marital status, level of education, area of residence, occupation and highly active antiretroviral therapy status (HAART) were recorded.

### Target condition

The infection (latent) status being targeted by the three tests (Xpert Ultra, SSM and LAM) constitutes a respiratory or urine sample carrying either the live MTB, its DNA or cell wall LAM antigen at any concentration.

### Sputum Smear Microscopy (SSM)

This was performed as per standard guidelines(7). Briefly, a sputum smear covering 2cm by 1 cm on the centre of the slide was prepared immediately after receipt of the sample. The smears were then stained with the primary stain 1% Carbol fuchsin for a period not exceeding 10 minutes. Decolourisation was then performed by flooding the slides with 25% sulphuric acid for 3 minutes and subsequently the counterstain 0.1% methylene blue was added for 1 minute. Slides were rinsed before every staining stage and excess water drained by gently tilting each slide. The slides were thereafter examined under a light microscope at 100x magnification. *Mycobacterium* quantification was achieved by counting the acid fast bacteria (AFB) as per the International Union Against TB and Lung Disease recommended grading of sputum smear microscopy(7). For analytical purposes, AFB counts above zero constituted a positive result; otherwise negative.

### Gene Xpert Mycobacterium/ Rifampicin Ultra (Xpert Ultra)

This was carried out as described elsewhere(26). Briefly, a sample reagent (a constitution of sodium hydroxide and Isopropanol) was added into a sputum sample in a ratio of 2:1 for liquefaction and inactivation of the sputum. The mixture was then vigorously shaken and incubated at room temperature for 15 minutes. The liquefied specimen was afterwards pipetted into the test cartridge (Cat. No. 42722, Cepheid). The pre-labelled test cartridge was loaded into the MTB/RIF test platform (Cepheid) where automated testing commenced. The semi-nested real time amplification and detection occurred in an integrated reaction tube and the results were printed. Results were interpreted as positive when MTB was detected.

### Lateral Flow Urine Lipoarabinomannan (LAM)

Alere Determine^TM^ TB LAM antigen kit (Cat No. 238237, Abbott Diagnostics Scarborough) was used to test for the presence of TB LAM antigen in collected urine samples. This was executed as per the instructions in the Alere Determine^TM^ TB LAM antigen manual (27). A total of 60ul of urine was added into the strip sample well. In order to allow for the urine to flow towards both the test and control windows, the strip was incubated at room temperature for 25 minutes. A positive result was denoted by the appearance of two colour bands; one band on the control line being indicative of a negative result.

### Statistical analysis

A Bayesian latent class model fitted in OpenBUGS v3.2.2(28) but called from R software through the ‘BRugs’ package(29) was used to estimate the TB prevalence, Se and Sp of the three tests together with their predictive values. The guidelines for standards for repo rting diagnostic accuracy studies that use BLCMs were used to inform the analysis and reporting of the results (30). The model code is available as underlying data(31).

Three assumptions are necessary when fitting a BLCM(32). Firstly, the tests should be conditionally independent given the TB infection status. This was met since microscopy targets the live bacterium, Xpert Ultra amplifies and detects the *Mycobacterium* DNA and TB LAM detects the *Mycobacterium* cell wall LAM antigen. Hence, the probability of testing either positive or negative on one diagnostic tool did not depend on the outcome of the other tests. Secondly, the Se and Sp of the tests being evaluated should be constant across the subpopulations studied. With a single target population, as was the case here, the tests’ constancy premise was likely to be upheld. Thirdly, the target population should consist of two or more subpopulations with different prevalences. In situations where only a single population exists, at least three tests are necessary to provide sufficient degrees of freedom to achieve model identifiability (30)

A multinomial distribution was assumed for the counts (𝑂) of the different test combinations (e. g. +, +, +) and took the form below:

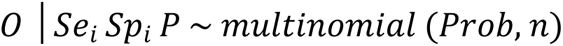

Where 𝑆𝑒_𝑖_ and 𝑆𝑝_𝑖_ reflect the specific test characteristics of the individual tests (*i* = 1, 2, 3), 𝑃 denotes the prevalence for the singular population, 𝑃𝑟𝑜𝑏 is a vector of probabilities of observing different test combinations and 𝑛 denotes the sample size of the study population. As an illustration, the probability of an individual testing positive on all three tests is given by:

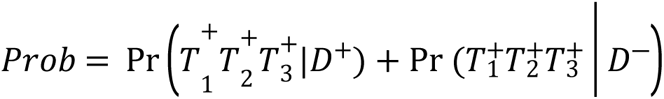

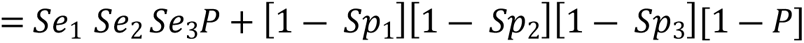

With three tests, a total of seven degrees of freedom was sufficient to estimate the required seven parameters (the Se and Sp of the three tests together with the single population prevalence). Since there was no reliable prior information for any of the tests, uninformative priors (𝑏𝑒𝑡𝑎 (1, 1)) were used as initial parameters for the model.

The positive predictive value (PPV) and negative predictive value (NPV) of the test (*i*) were estimated as follows:

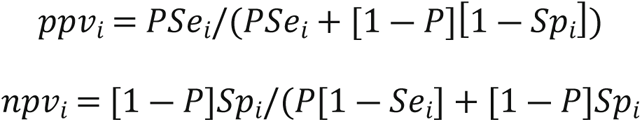

Two Markov Chain Monte Carlo chains each having different values were used to initialise the Bayesian model. Convergence of the chains was then evaluated via the time series plots of selected variables and the Gelman-Rubin diagnostic plots. The posterior distributions of the test estimates (Se, Sp and the predictive values) and the population prevalence were reported as the median and the associated 95% posterior credible intervals (PCI). The Youden index which is a measure of a test’s overall diagnostic ability was also computed as : 𝑆𝑒 + 𝑆𝑝 ― 1 (33).

## Results

Of the 190 participants, two were unable to provide all the required samples and were thus excluded from the analysis. Another five were excluded on the basis of their Xpert Ultra results being invalid.

The sociodemographic characteristics and information on HAART status is as shown in Table 1. The median age of the participants was 43 years (Range: 19 – 82 yrs). The majority of participants were female (64.0%, n=117), married (60.7%, n=111), with only 2.2% (n=4) lacking a formal education. A substantial percentage of the participants was on HAART (97.5%, n=178).

**Table 1:** Summary statistics on the sociodemographic characteristics and information on HAART status for HIV patients presenting to a primary health facility in an informal settlement, Nairobi County, Kenya. (n=183)

| Variable | Values | Median | Range | Frequency n (%) |
| --- | --- | --- | --- | --- |
| **Age** |  |  |  |  |
|  |  | 43 | 19-82 |  |
| **Weight** |  |  |  |  |
|  |  | 60 | 40-101 |  |
| **Sex** |  |  |  |  |
|  | Male |  |  | 66(36.1) |
|  | Female |  |  | 117(64.0) |
| **Marital status** |  |  |  |  |
|  | Married |  |  | 111(60.7) |
|  | Single |  |  | 50(27.3) |
|  | Separated/ divorced |  |  | 18(9.8) |
|  | Widowed |  |  | 4(2.2) |
| **Level of education** |  |  |  |  |
|  | None |  |  | 4(2.2) |
|  | Primary |  |  | 91(49.7) |
|  | Secondary |  |  | 69(37.7) |
|  | Tertiary |  |  | 19(10.4) |
| **Area of residence** |  |  |  |  |
|  | Formal |  |  | 60(32.8) |
|  | Informal |  |  | 123(67.2) |
| **Occupation** |  |  |  |  |
|  | Employed |  |  | 67(36.6) |
|  | Unemployed |  |  | 116(63.4) |
| **HAART** |  |  |  |  |
|  | Naïve |  |  | 5(2.7) |
|  | Experienced |  |  | 178(97.3) |

HAART; Highly Active Antiretroviral Therapy

The cross-tabulated counts of the tests’ outcomes are displayed in Table 2.

**Table 2:** Cross-classified results for LAM, Xpert Ultra and SSM tests for diagnosis of TB among PLWHIV presenting to the Kibra Community Health Centre CCC clinic, Nairobi County, Kenya. (n=183)

Cross-tabulated counts of the three test outcomes
| Test outcomes combinations (LAM, XPERT ULTRA & SSM) |  |  |  |  |  |  |  |  |  |
| --- | --- | --- | --- | --- | --- | --- | --- | --- | --- |
| Stratum |  |  |  |  |  |  |  |  | Total (%) |
|  | +++ | ++- | + - + | - + + | + - - | - + - | - - + | - - - |  |
| Single population | 1 | 1 | 0 | 3 | 21 | 8 | 0 | 149 | 183 (100%) |

LAM; Lipoaribomannan, SSM; Sputum Smear Microscopy

The estimates of the Se and Sp of the three tests, along with their respective predictive values and the prevalence of MTB infection are shown in Table 3. The Xpert Ultra assay registered a higher Se (85.0; 95% PCI [41.4 – 99.4]) compared to LAM (26.8; 95% PCI [4.7 – 67.6]) and SSM (56.7 [16.4 – 97.4]). However, SSM displayed the highest Sp (99.6; 95% PCI [97.7 – 100.0]). As per the Youden indices, Xpert Ultra yielded the highest overall combination of Se and Sp at 80.8% (95% PCI [37.0 – 96.5]). On predictive values, SSM recorded the highest PPV at 84.5% (95% PCI [38.4 – 99.4]). Nonetheless, all the tests exhibited noticeably high NPVs (>96%). The true prevalence of MTB infection in this HIV-infected outpatient population was 4.1% (95% PCI [1.2 – 11.2]).

**Table 3.**
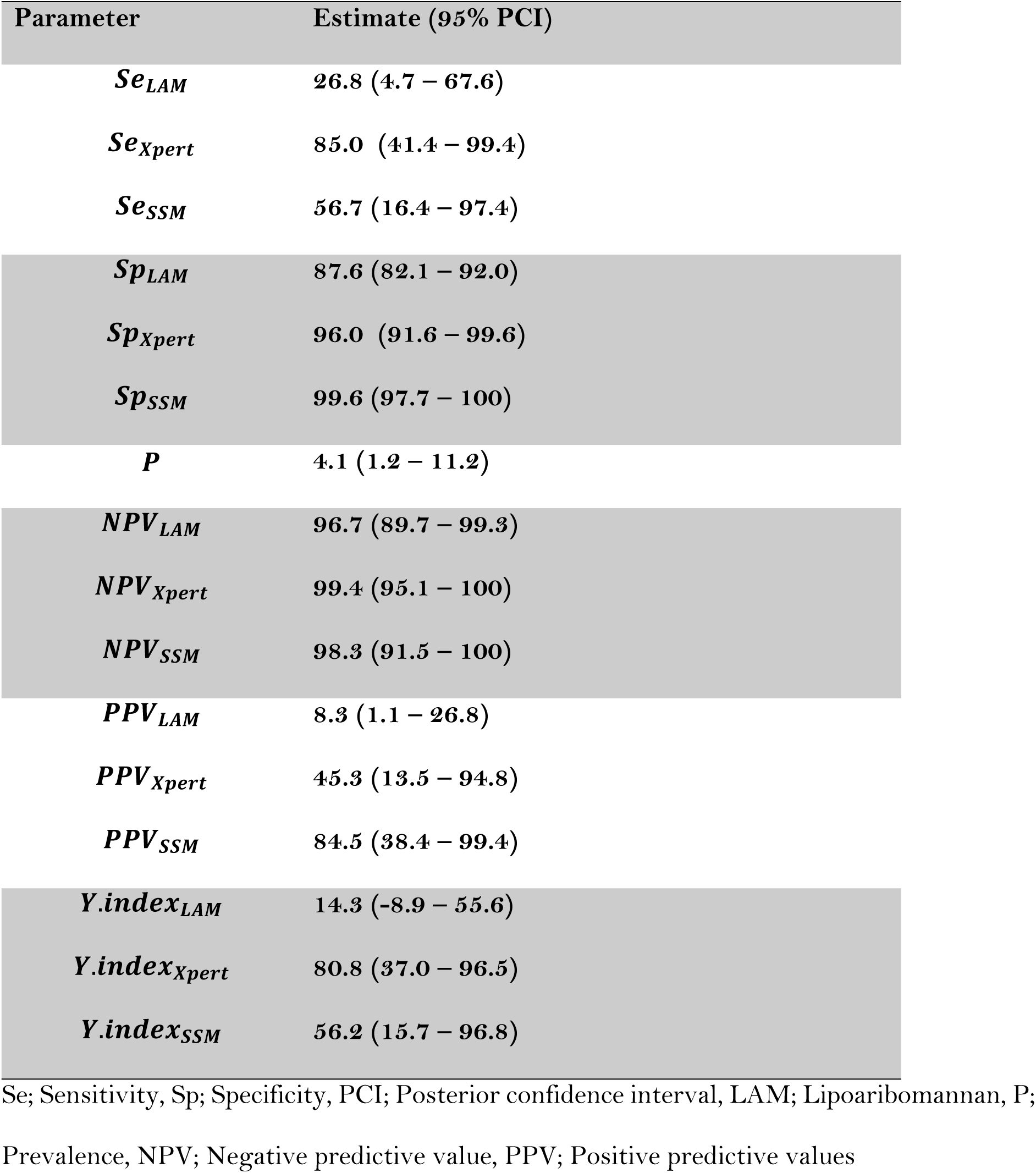
Estimates of prevalence, sensitivity, and specificity of LAM, Gene Xpert Ultra, and SSM tests for MTB infection among HIV patients as well as their corresponding predictive values and Youden indices.

## Discussion

This study has utilized Bayesian latent class analysis for the estimation of the accuracy of SSM, Xpert Ultra and LAM tests (together with their corresponding predictive values) for the diagnosis of TB infection among PLWHIV. This is a key strength in this study since BLCM allows for the estimation of index tests’ characteristics devoid of classification errors typically inherent in traditional evaluation studies employing the use of imperfect reference standards(21). Accordingly, the estimates obtained from this study can be considered generalisable to settings with similar MTB burden among PLWHIV.

On Se, Xpert Ultra assay recorded the highest Se of the three tests. This result is in agreement with previous reports that demonstrated the superiority of Xpert Ultra Se over other tests in TB screening and diagnosis among PLWHIV in outpatient settings. In particular, in a systematic review involving studies conducted in high TB prevalence settings among PLWHIV, the Xpert Ultra test Se ranged between 81% - 90%(20). The superiority of the test’s Se owes to its low detection limit of tubercle bacilli (16 CFU/ ml of sputum)(34) compared to SSM whose threshold is 5,000 - 10,000 CFU/ ml of sputum (35) and LAM at ∼ 2000 CFU / ml (36, 37).

The LAM test exhibited the lowest Se in the diagnosis of TB in the present study. This performance is comparable with findings from studies carried out in other outpatient settings. For instance, among existing, ART-naïve and newly diagnosed HIV patients presenting to outpatient facilities in LMICs, LAM’s Se was found to range between 25% - 30% (17, 36, 38, 39). Notably, an inverse relationship has been demonstrated, with LAM’s test Se increasing with reducing CD4 counts among critically ill HIV patients(17, 40). LAM Se was observed to be at its highest (≥65%) among patients with CD4 counts <50 cells/ul and Se of35%-55% for patients with CD4 counts between100 and 200 cells/ul respectively (17, 40). Increased uptake of ARV therapy in the present study setting may have contributed to improved CD4 counts in the patient population resulting in a low Se of the LAM test - underscoring its limited utility in this setting. SSM registered a test Se of 56.7% which falls within the 18% - 94.2% range observed in other studies(41, 42). The wide variability in performance is attributable to the number of CFUs available in a sample, with Se increasing with higher CFUs. Moreover, the test Se is also highly dependent on user expertise and other technical and operational factors (42).

As regards Sp, SSM exhibited the highest which is consistent with findings from studies carried out in outpatient settings among PLWHIV(43, 44). The Sp displayed by LAM is also corroborated by findings from other studies carried out in similar low-resource settings where specificities ranging between >90%-100% were registered (36, 38, 45, 46). Xpert Ultra Sp is in agreement with specificities found in other similar settings where the test Sp was reported to range between 78%- 96% (20, 47).

False negative and false positive results compromise Se and Sp estimates respectively. As for LAM and SSM, false negative results are more likely to occur in patients with bacterial load below the tests detection limits(48). Additionally, LAM is more likely to give false negative results in patients who have already received anti-TB treatment(49). False positive results by LAM have been demonstrated in samples contaminated with non-Mycobacterial pathogens such as *Nocardia* and *Candida* spp. that similarly exhibit LAM-like glycoprotein antigen(46). SSM is also unable to distinguish MTB from other smear-positive *Mycobacteria* spp.(50). False positives by Xpert Ultra could be attributable to detection of MTB DNA in patients recently cured of TB (44).

Notably, in this population, the three tests’ sensitivities displayed wide credible intervals (signifying low precision of the estimates) since there were few diseased individuals. In contrast, the tests’ Sps demonstrated high precision owing to the significant number of truly non-diseased individuals. Overall, in this low-prevalence TB setting, Xpert Ultra affords good promise for informing treatment and surveillance for TB among PLWHIV.

With low MTB prevalence (4.1%), generally low PPVs but high NPVs would be anticipated for the three tests. These estimates signify a stronger confidence in a negative than a positive test result. Consequently, in this low-prevalence setting where the probability of false positives is highly contemplated, a multiple testing strategy with serial interpretation of the test results may be necessary in order to raise the confidence in a positive test result. This could entail an initial screening with the more sensitive Xpert Ultra test, with any resulting positives followed up with the more specific SSM test. This approach should assure that false positive individuals are not unnecessarily subjected to protracted therapy for TB.

## Conclusions

The Xpert Ultra assay registered the highest Se compared to SSM and LAM. However, SSM registered the highest Sp and thus PPV estimate. Nevertheless, the three tests recorded similar NPVs. Owing to the low prevalence of TB in the study setting, an optimal testing approach could entail an initial screening with the more sensitive Xpert Ultra assay, with any resultant positives re-tested with the more specific SSM test – a serial testing strategy - to bolster the overall PPV of a TB testing and surveillance programme.

## Data Availability

All underlying data files are available from the Havard dataverse database https://doi.org/10.7910/DVN/PYRPCQ

## Acknowledgments

The authors sincerely thank the study participants, the administration and staff at Kibra Community Health Centre for making it possible to realise this research work.

## Supporting information

**S1 File**. **Harvard Dataverse: Replication Data for: Performance of three diagnostic tests for tuberculosis among HIV patients presenting to a primary care facility in an informal settlement in Nairobi County, Kenya**. https://doi.org/10.7910/DVN/PYRPCQ(31)

